# Blinded study: prospectively defined high frequency oscillations predict seizure outcome in individual patients

**DOI:** 10.1101/2020.12.24.20248799

**Authors:** V Dimakopoulos, P Mégevand, E Boran, S Momjian, M Seeck, S Vulliémoz, J Sarnthein

## Abstract

**Background:** Interictal high frequency oscillations (HFO) are discussed as biomarkers for epileptogenic brain tissue that should be resected in epilepsy surgery to achieve seizure freedom. The prospective classification of tissue sampled by individual electrode contacts remains a challenge. We have developed an automated, prospective definition of clinically relevant HFO in intracranial EEG (iEEG) from MNI Montreal and tested it in iEEG from Zurich. We here validate the algorithm on iEEG recorded in an independent epilepsy center so that HFO analysis was blinded to seizure outcome.

**Methods:** We selected consecutive patients from Geneva University Hospitals who underwent resective epilepsy surgery with postsurgical follow-up > 12 months. We analyzed long-term iEEG recordings during non-rapid eye movement (NREM) sleep that we segmented into intervals of 5 min. HFOs were defined in the ripple (80-250 Hz) and the fast ripple (FR, 250-500 Hz) frequency band. Contacts with the highest rate of ripples co-occurring with FR (FRandR) designated the HFO area. If the HFO area was not fully resected and the patient suffered from recurrent seizures (ILAE 2-6), this was classified as a true positive (TP) prediction.

**Results:** We included iEEG recordings from 16 patients (median age 32 y, range [18-53]) with stereotactic depth electrodes and/or with subdural electrode grids (median follow-up 27 mo, range [12-55]). The HFO area had high test-retest reliability across intervals (median dwell time 95%). We excluded two patients with dwell time < 50% from further analysis.

The HFO area was fully included in the resected volume in 2/4 patients who achieved postoperative seizure freedom (ILAE 1, specificity 50%) and was not fully included in 9/10 patients with recurrent seizures (ILAE > 1, sensitivity 90%), leading to an accuracy of 79%.

**Conclusions:** We validated the automated procedure to delineate the clinical relevant HFO area in individual patients of an independently recorded dataset and achieved the same good accuracy as in our previous studies.

**Significance:** The reproducibility of our results across datasets is promising for a multicienter study testing the clinical application of HFO detection to guide epilepsy surgery.

## Introduction

Drug-resistant focal epilepsy is a common condition. In selected patients, surgical resection of the epileptogenic zone (EZ) is the treatment of choice and may eliminate occurrence of seizures completely (1). The EZ is defined as the brain area whose resection leads to freedom from seizures (2). Preoperative diagnostic workup may involve recording of the intracranial EEG (iEEG) to determine the seizure onset zone (SOZ) as an estimate for the EZ. Since seizures are usually rare events during the limited duration of iEEG recording, it would be advantageous to determine the EZ during the interictal period. In this approach, the traditional analysis of interictal epileptiform discharges has a high sensitivity but low specificity as an interictal marker of epileptogenic tissue (3), which may be improved by more advanced analysis (4).

As a further marker, high frequency oscillations (HFOs) may have the potential to be a clinical asset for delineating epileptogenic brain areas and identifying successful surgical treatments (3, 5-10). These oscillatory events can be found in the frequency range between 80–500 Hz. HFOs are sub-classified in ripples (80-250 Hz) and fast-ripples (FRs, 250-500 Hz). Interictal HFOs that mostly occur during NREM sleep have proven more specific than interictal spikes in localizing the SOZ or ‘predicting’ seizure outcome (11-16). While many studies present HFO rates in relation to SOZ electrodes (17), only in a subset of studies was the resection of interictal HFOs, marked prospectively, proposed for the ‘prediction’ of post-surgical seizure freedom (7, 14, 15, 18-20).

Investigations in the clinical relevance of the HFOs have been facilitated by automated or semi-automated detection algorithms (20). Here we apply a fully automated definition of HFOs, which we previously optimized on visual markings in a dataset of the Montreal Neurological Institute (21) and then validated on independently recorded data from Zurich (7). We thus provide a prospective definition of a clinically relevant HFO where we focus on ripples (80-250 Hz) co-occurring with FRs (250-500 Hz), which we call FRandR. For each recording interval, we define as HFO area the electrode contacts with the HFO rate exceeding the 95% percentile of the HFO distribution. We then test the reliability of the HFO area over multiple recording intervals.

In the present study, we applied this algorithm to iEEG recorded in an independent epilepsy center (HUG, Hôpitaux Universitaires de Genève, Switzerland). The analysis was blind with respect to clinical outcome. We compared the HFO area with the resected brain volume (RV) and ‘predicted’ the seizure outcome in individual patients in order to evaluate the clinical relevance of HFO analysis.

## Materials and Methods

### Patients

We included patients with drug-resistant focal epilepsy who 1) underwent invasive EEG recordings with subdural and/or depth electrodes as part of their presurgical evaluation at HUG between 2015 and 2019, 2) underwent resective surgery and 3) were followed for at least one year after surgery. The decision for resective surgery was based on non-invasive investigations as well as on intracranial investigations (3). The results of HFO analysis were not used for surgical planning. The postsurgical seizure outcome was determined by follow-up visits and classified according to the International League Against Epilepsy (ILAE).

### Electrode types and implantation sites

Subdural grid electrodes as well as depth electrodes were placed according to the findings of the non-invasive presurgical evaluation. In 15 patients, depth electrodes (varying electrode configurations, ADTech®, www.adtechmedical.com, and Dixi Medical, www.diximedical.com) were implanted stereotactically. In Patient 10, additional subdural electrodes were inserted through a craniotomy. In Patient 16, subdural grid and strip electrodes (contact diameter 4 mm with 2.3 mm exposure, spacing between contact centers 10 mm, ADTech®) were placed after craniotomy. Pre-implantation MR and post-implantation CT images were used to locate each electrode contact anatomically using the intracranial electrode localization and visualization (iELVis) toolbox (**Figure 1A**) (22).

**Figure 1:**
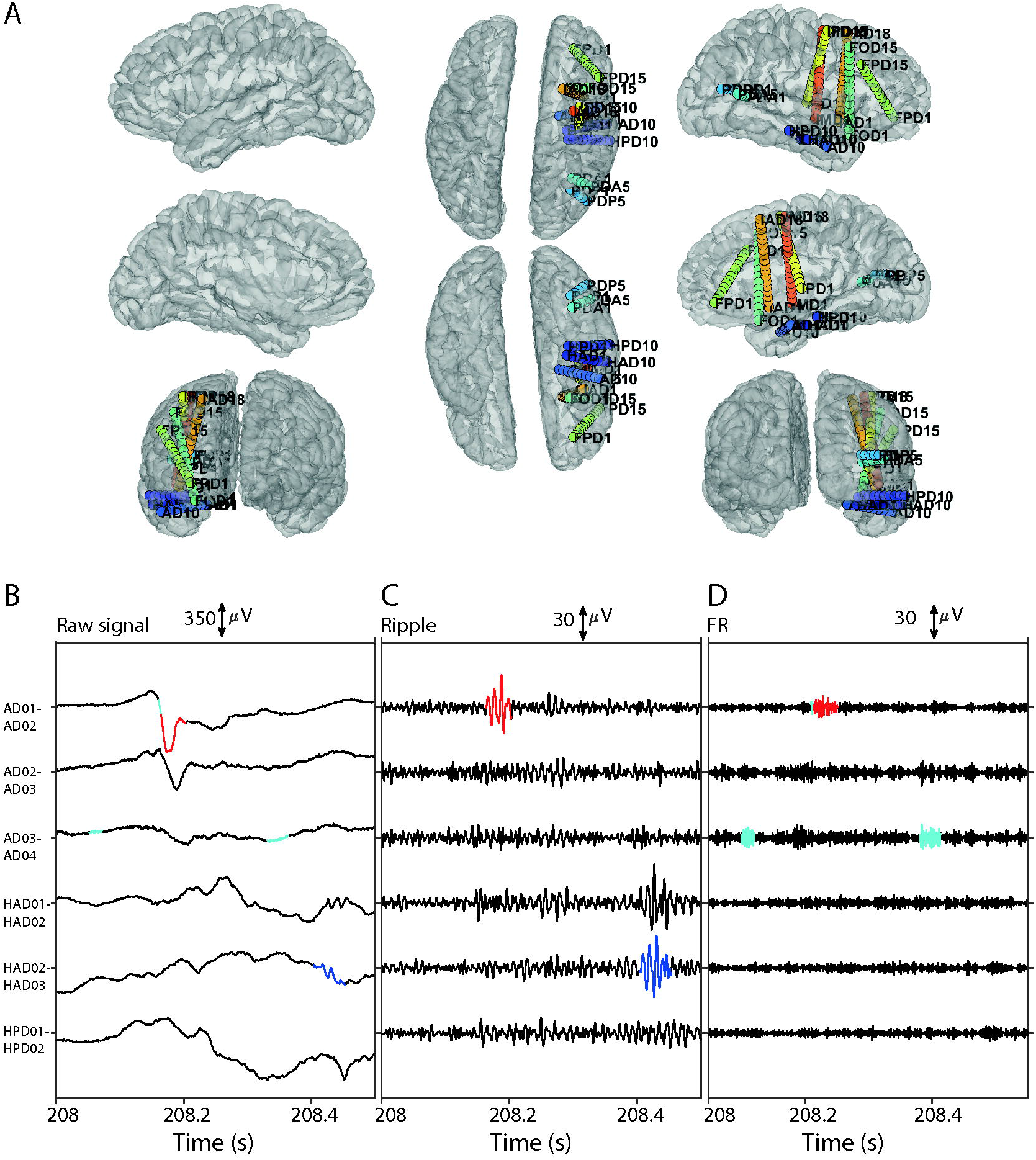
Automated HFO analysis in Patient 11. (A) Locations of the iEEG depth electrodes. FPD □=□orbitofrontal posterior right; FOD □=□orbitofrontal right; AD □=□amygdala right; HAD□=□ hippocampus anterior right; HPD□=□ hippocampus posterior right; PDP □=□parietal posterior right; PDA □=□parietal anterior right; IAD □=□insula anterior right; IPD□=□insula posterior right; IMD =□insula middle right; CPD = cingulate posterior right (B, C, D) HFO detection: example of a ripple co-occurring with an FR (FRandR). (B) Wideband iEEG (C) iEEG filtered in the ripple band [80 250] Hz (D) iEEG filtered in the fast ripple band [250 500] Hz. The HFO detection is highly specific: while several ripples and FRs were detected, only one FRandR was selected as a clinically relevant HFO. Ripple (R): blue; Fast ripple (FR): cyan; FRandR: red

### Data preprocessing

We selected data that were recorded during nights while the patient was in NREM sleep. We then divided the data into 5-minute intervals for further analysis. The data was resampled from 2048 Hz to 2000 Hz using the polyphaser anti-aliasing filter in Matlab. The iEEG was recorded against a common subcutaneous reference placed around the vertex and then transformed to bipolar channels.

We identified channels from sensorimotor and occipital brain regions using iELVis, because these are thought to exhibit large numbers of physiological HFO (23).

### Prospective definition of HFOs

HFOs were defined prospectively by the automated detector, which we had previously trained and validated to detect visually marked events in datasets from the Montreal Neurological Institute (21) and which was then validated in an independent dataset from Zurich (7).

In brief, the detector incorporates information from both time and frequency domain and operates in two stages. In the first stage – baseline detection – the Stockwell transform identified high entropy segments with low oscillatory activity. The values of the envelope of the signal at these high entropy segments define the baseline. The second stage – HFO detection – was conducted separately for ripples (band-pass 80– 240□Hz, stopband 70□Hz and 250□Hz, FIR equiripple filter with stopband attenuation 60□dB) and FRs (band-pass 250– 490□Hz, stopband 240□Hz and 500□Hz). Events with the envelope of the filtered signal exceeding the amplitude threshold for at least 20□ms/10□ms were labelled as ripples/FR **(Figure 1B, C)**. The algorithm then identified FRs overlapping with a ripple, which we defined as a third type of HFO: FR co-occurring with ripples (FRandR, **Figure 1D**). There was no manual rejection of events in this fully automated algorithm.

### Definition of the HFO area by rate thresholding

In each recording interval and each patient, we computed the HFO rate by dividing the HFO count per channel by the duration of the epoch in minutes. We then analyzed the spatial distribution of HFO rates across channels. The ensemble of those channels whose rates exceeded the rate threshold (95^th^ percentile of the HFO rate distribution) was defined as the HFO area **(Figure 2A)**.

**Figure 2:**
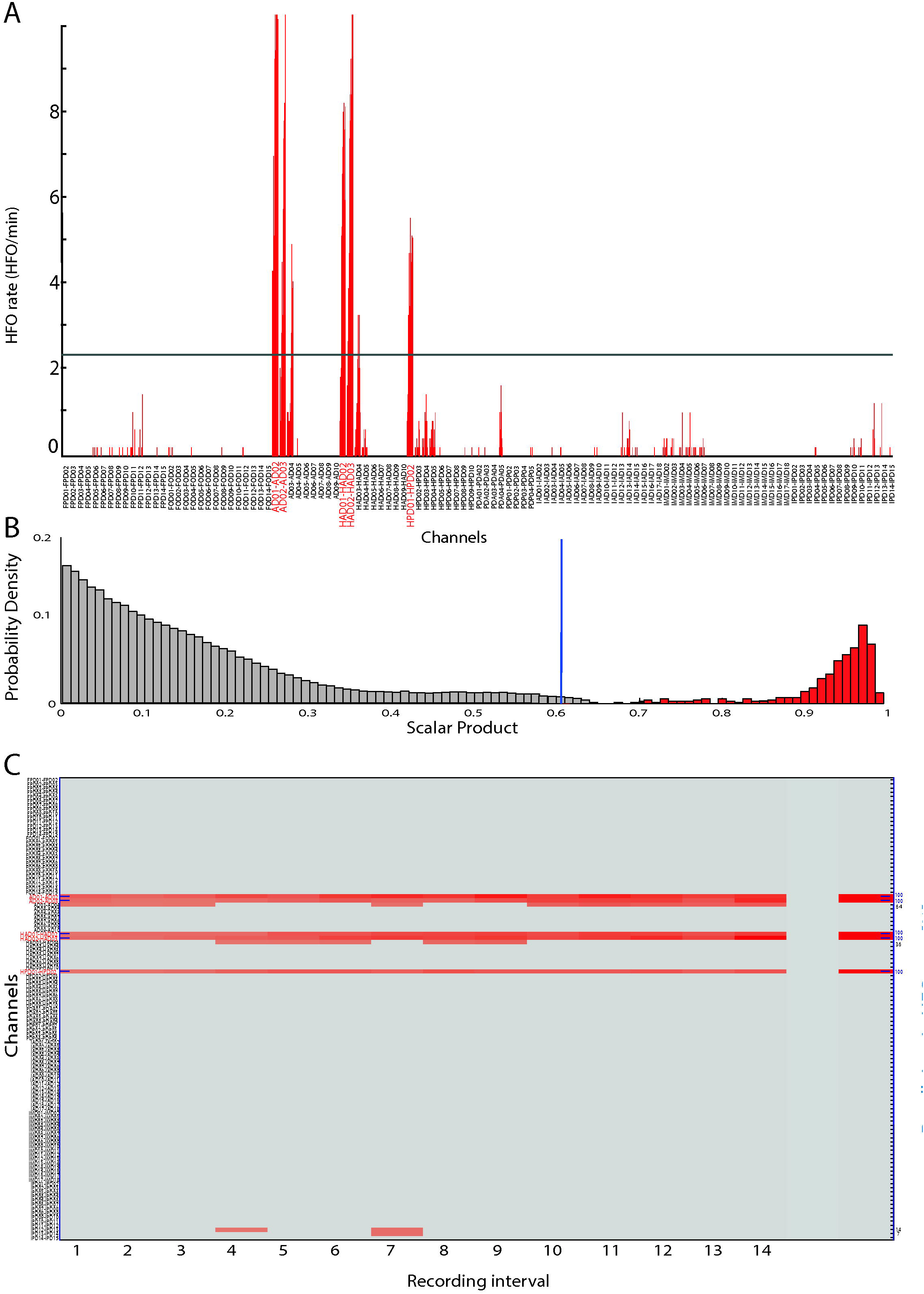
Test-Retest analysis of HFO rates for Patient 11. (A) HFO rate (FRandR, co-occurring ripple and fast ripple, HFO/min) for each interval (red vertical bar). Channels with rates that exceed the 95^th^ percentile (black line) are candidates to be included in the HFO area (rate thresholding). (B) The anatomical distribution of HFOs is not random. The true distribution of the normalized scalar product of HFO rates for each pair of intervals (red). The random distribution of the normalized scalar product of HFO rates obtained by permutation analysis (gray, 10000 permutations). The 97.5^th^ percentile of the random permutation (vertical blue line) serves as the significance threshold. 100% of the true distribution exceed the significance threshold; therefore the anatomical distribution of HFOs is not random. (C) Reproducibility of the HFO area over recording intervals. Red bars denote channels where the HFO rate exceeds the 95^th^ percentile in that interval. Channels from the tip of recording electrodes AD, HAD and HPD have red bars for a dwell time = 100% of the recording intervals. AD = amygdala right; HAD = hippocampus anterior right; HPD = hippocampus posterior right

### Reliability of the spatial distribution of the HFO area

We then tested whether the HFO area was simply a product of chance. We selected each interval pair and computed the normalized scalar product of the spatial distribution of the HFO rates (**Figure 2B**). The scalar product is 1 for highly overlapping spatial distributions of HFO rate and lower otherwise. To test the magnitude of the true scalar product against chance, we constructed a distribution of scalar products by randomly permuting (N□=□10000) the order of channels for each interval. The true value of the scalar product was considered statistically significant if it exceeded the 97.5% percentile of the distribution We report the percentage of interval pairs where the scalar product was significant (**Table 1**). Finally, we quantified the test-retest reliability of the HFO area over the ensemble of recording intervals by counting the percentage of intervals that each channel spent in the HFO area (dwell time, **Figure 2C**). Patients with median dwell time < 50% were excluded from further analysis (7).

**Table 1.** Patient Characteristics. Abbreviations: HS = hippocampal sclerosis; FCD = focal cortical dysplasia; WM = white matter; ILAE□=□International League Against Epilepsy; TP = True Positive; TN = True Negative; FP = False Positive; FN = False Negative.

| i | Histology | Electrode type | Contacts | Intervals | Test-retest intervals (%) | Median dwell time (%) | Outcome (ILAE scale) | Follow-up (months) | Prediction ILAE >1 |
| --- | --- | --- | --- | --- | --- | --- | --- | --- | --- |
| 1 | No HS, no dysplasia | depth | 42 | 25 | 100 | 89 | 1 | 48 | TN |
| 2 | No abnormality | depth | 70 | 21 | 63 | 74 | 1 | 34 | TN |
| 3 | FCD type IIb | depth | 70 | 10 | 90 | 83 | 1 | 36 | FP |
| 4 | HS, WM ectopic neurons | depth | 177 | 12 | 95 | 100 | 1 | 18 | FP |
| 5 | Hippocampal gliosis | depth | 76 | 17 | 84 | 75 | 4 | 32 | TP |
| 6 | WM ectopic neurons | depth | 77 | 8 | 100 | 100 | 5 | 12 | TP |
| 7 | Cortical gliosis | subdural | 81 | 13 | 88 | 79 | 4 | 18 | TP |
| 8 | WM ectopic neurons | depth, subdural | 82 | 21 | 80 | 94 | 3 | 30 | TP |
| 9 | WM ectopic neurons | depth | 123 | 14 | 96 | 89 | 3 | 55 | TP |
| 10 | FCD type IIa | depth | 126 | 22 | 100 | 89 | 4 | 24 | TP |
| 11 | WM ectopic neurons | depth | 129 | 17 | 100 | 100 | 3 | 21 | TP |
| 12 | HS | depth | 162 | 26 | 98 | 88 | 3 | 21 | TP |
| 13 | WM ectopic neurons | depth | 197 | 15 | 100 | 100 | 4 | 18 | TP |
| 14 | No HS, no dysplasia | depth | 106 | 17 | 98 | 100 | 5 | 30 | FN |
| 15 | No dysplasia | subdural | 74 | 27 | 43 | 45 | 1 | 16 | -- |
| 16 | WM ectopic neurons | subdural | 112 | 16 | 52 | 46 | 3 | 45 | -- |

**Table 2.** HFO area and seizure freedom. Resection of the HFO area has intermediate specificity in predicting the seizure freedom and is highly sensitive in predicting seizure recurrence. Abbreviations: TP□=□True Positive; TN□=□True Negative; FP□=□False Positive; FN□=□False Negative; N□=□number of patients; PPV□=□Positive Predictive Value; NPV□=□Negative Predictive Value;

|  |  |  |
| --- | --- | --- |
| Sensitivity | TP/(TP + FN) | 90% |
| Specificity | TN/(TN + FP) | 50% |
| Positive Predictive Value PPV | TP/(TP + FP) | 82% |
| Negative Predictive Value NPV | TN/(TN + FN) | 67% |
| Accuracy | (TP + TN)/N | 79% |

### Clinical validation of HFO against seizure outcome

Automated HFO detection and analysis were blind to clinical information. We evaluated whether the HFO area was included in the resected volume (RV) to quantify the predictive value of the HFO area with respect to seizure outcome. The electrode positions in the RV were determined from post-resection MR coregistered to pre-implantation MR scans (22). Electrodes landing on the border of the resection were deemed to be part of the RV. We defined as true positive (TP) a patient where the HFO area was not fully located within the RV, i.e. at least one channel of the HFO area was not resected, and the patient suffered from recurrent seizures (ILAE 2–6). We defined as false positive (FP) a patient where the HFO area was not fully located inside the RV but who achieved seizure freedom (ILAE 1). We defined as false negative (FN) a patient where the HFO area was fully located within the RV but who suffered from recurrent seizures. We defined as true negative (TN) a patient where the HFO area was fully located inside the RV and who became seizure free. The positive predictive value was calculated as PPV□=□TP/(TP□+□FP), negative predictive value as NPV□=□TN/(TN□+□FN), sensitivity□=□TP/(TP□+□FN), specificity□=□TN/(TN□+□FP), and accuracy□=□(TP□+□TN)/N.

### Statistics

We used the Wilcoxon rank-sum test to compare distributions. We compared percentages with the chi-square test. We estimated the 95% confidence intervals (CI) of proportions by the binomial method. All statistical analyses were performed in Matlab. Statistical significance was established at p<0.05.

## Results

### Patients and seizure outcome

We included 16 patients in the study that fulfilled the inclusion criteria (**Table 1**). Complete seizure freedom (ILAE 1) was achieved in 5 patients (seizure-free rate 31%), while 11 patients suffered from seizure recurrence (ILAE 2 – 6). 10 of 16 patients (63%) experienced significant reduction in their seizure burden (ILAE 1-3). Two patients were excluded from further analysis because the median dwell time of channels in the HFO area was <50%, which was due to persistent artifacts in the iEEG in the HFO frequency range. In the remaining patient group (N = 14), the mean follow-up for good outcome (34 ± 12 mo) was longer than for poor outcome (26 ± 12 mo) but the two distributions are indistinguishable (p□=□0.22 Wilcoxon rank-sum test).

### HFO detection and test-retest reliability of the HFO area

The HFO detection algorithm was applied to the recordings of all 16 patients. The number of nights and intervals varied across patients (Table 1). Over the group of patients we identified ripples (median amplitude 17.4 μVpp, interquartile range 9.8 μVpp) and FR (median amplitude 10.6 μVpp, interquartile range 3.7 μVpp). We used the co-occurrence of ripples and FR (FRandR) to determine the HFO area in each patient. The channels in sensorimotor and occipital brain regions were never in the HFO area. (**Supplementary Table 1**).

### The HFO area predicts seizure outcome in individual patients

For each of the 14 patients, we evaluate whether the HFO area was fully or partly resected. The HFO area was fully resected in 2 patients who achieved seizure freedom (TN). The HFO area was not fully resected in 9 patients who did not achieve seizure freedom (TP). The HFO area was not fully resected in 2 patients who nevertheless achieved seizure freedom (FP). The HFO area was fully resected in 1 patient who did not achieve seizure freedom (FN).

From these values we obtain specificity = 50% CI [67 93%], sensitivity = 90% CI [55 99%], NPV = 67%, CI [9 99%], PPV = 89% CI [48 97%], and accuracy = 79% CI [49 95%]. The low specificity is related to the small number of correctly predicted seizure-free patients (TN = 2 / 4 patients with ILAE 1). The high sensitivity is explained by the high number of patients where the recurrence of seizures was correctly predicted (TP = 9 / 10 patients with ILAE > 2).

Of the patients where recurrent seizures were correctly predicted (TP = 9), the HFO area was not fully resected in 4 patients (44%), and the HFO area was completely dissociated from the RV in 5 patients (56%) **(Supplementary Table 1)**.

When compared to the SOZ, the HFO area matched the SOZ completely in 7/14 and partially in 2/14 patients **(Supplementary Table 1)**.

Both in our analysis and current surgical planning, the FN case may stem from the limited coverage of the implanted electrodes. The very high sensitivity (90%) and PPV (89%) suggest that automated HFO detection might have contributed to improved surgical planning, since 9 of the 10 patients in whom the HFO area was not fully resected suffered from recurrent seizures.

### Combining the Geneva cohort and the Zurich cohort

If we combine the Geneva cohort of this study (N = 14) and the Zurich cohort (N = 20) (7) that were analysed with the same HFO detector, we obtain specificity = 88% CI [63 98%], NPV = 79% CI [54 93%], sensitivity = 76% CI [47 92%], PPV = 87% CI [57 98%], accuracy = 82% CI [64 93%]. The predictionaccuracy for this combined cohort is associated with the surgical planning (seizure free rate 50% CI [33 67%], p = 0.001 chi-square test).

## Discussion

### Methodological considerations

The delineation and the clinical evaluation of the HFO area provided high sensitivity (90%) in predicting seizure recurrence: if the HFO area is not fully resected then seizure freedom ILAE 1 is not achieved. The high sensitivity could be associated with the capability of the HFO to generalize across individual patients. Contrary to a previous multicenter study (24), we were able to accurately correlate the seizure outcome with the HFO area. This discrepancy might be explained by the definition of a clinically relevant HFO; based on our previous study, we define the co-occurrence of ripples and fast ripples (FRandR) as biomarker for EZ, which has been proven more specific than ripples or fast ripples (7).

The detection of HFOs can be challenging because of their low signal-to-noise ratio and artifacts in the iEEG. In meeting this challenge, our HFO detector was designed for HFO detection during NREM sleep (7, 21). The automated analysis pipeline results in a prospective definition of the HFO area. Distinct from other studies that consider only visual markings, we use here the test-retest reliability of the spatial distribution of the HFO. The reproducibility of the HFO area, within and over the data intervals, can be explained by the high internal consistency of the HFO rates over the intervals and provides a reliable biomarker for the epileptogenic zone. In two patients (15, 16), the test-retest reliability was low (dwell time < 50%); visual inspection indeed confirmed a large number of artifacts in these recordings, which caused spurious HFO detection. In this way, the test-retest approach helps to make the HFO analysis pipeline more reliable.

Interestingly, there were HFOs both inside and outside of the HFO area. This may have several reasons: Epilepsy is a network disease and HFOs appear at distributed locations of the network. The limited time of iEEG recording during a few days may point to an HFO area that might not reflect the EZ. Our prospective definition of HFOs was validated against the resection of the EZ. Our prospective definition of a clinically relevant HFO focuses on the co-occurrence of a ripple and a fast ripple (FRandR). It thereby ignores the traditional distinction between ripples (80-250 Hz) and FR (250-500 Hz) (25). While the rate of FRandR is obviously lower than the rates of ripples and FR separately, this definition may still be highly sensitive but not very specific, e.g. events might be labelled as epileptic FRandR, which in fact they are not. In view of these considerations, we had introduced the 95% rate threshold to label a channel as member of the HFO area and the consistency of this labeling over time to gauge the reliability of the HFO area (7).

In our cohort, the rate of post-operative seizure freedom was relatively low (31%), although a majority of patients (63%) experienced a significant reduction in their seizure burden. In our opinion, this reflects the clinical practice of the study center, where iEEG implantation is reserved to the most complex cases, including a large proportion of patients with extra-temporal lobe epilepsy and MRI-negative epilepsy (26).

### Clinical relevance and generalizability

In order for HFO analysis to obtain clinical relevance, HFOs must be validated against seizure outcome, should be defined prospectively, and should be tested on sufficient data (8). The results of our algorithm suggest that the information provided by prospectively defined HFOs could contribute to surgery planning in patients where the extent of surgical resection is difficult to define and can be adapted. It is in these patients where complementary electrophysiological markers such as HFO may be useful (5, 15, 27). Different from epileptic spikes, HFOs do not propagate, which is an advantage if the EZ needs to be localized precisely (9, 16). A growing number of studies relates the presence of HFOs to surgical outcome (20, 28). A prospective, automated definition of HFOs renders HFO analysis more generalizable (7, 18, 19). Further, the algorithm used here was previously trained and validated on datasets from two epilepsy centers (7, 21). Its good performance on an independent third dataset supports the generalizability and clinical relevance of HFO analysis.

## Conclusions

In a blinded study design, we validated the automated procedure to delineate the clinical relevant HFO area in individual patients of an independently recorded dataset. HFO analysis showed a very good sensitivity and PPV, i.e. if HFO remained outside the resection volume, it was more likely that the patient continued to seize. Together with an intermediate specificity and NPV, this achieved the same good accuracy as in our previous studies. The reproducibility of our results across datasets is promising for a multicenter study testing the clinical application of HFO detection to guide epilepsy surgery.

## Data Availability

Data will be published upon acceptance of the article.

## Acknowledgements

We acknowledge grants awarded by the Swiss National Science Foundation (SNSF 176222 to JS, SNSF 192749 to SV) and a SNSF Ambizione fellowship (PZ00P3_167836 to PM). The collaboration between the members of Université de Genève and Universität Zürich benefitted from joint seed money funding. The funders had no role in the design or analysis of the study.

## Data availability

All data needed to evaluate the conclusions in the paper are present in the paper.

The iEEG recordings and the electrode positions will be made publicly available upon acceptance of the article.

## Code availability

The code of the HFO detector is freely available at the repository https://github.com/ZurichNCH/Automatic-High-Frequency-Oscillation-Detector.

### Ethics statement

The study was approved by the research ethics committees (Cantonal ethics commissions of Zurich and of Geneva) and waived collection of patients’ written informed consent 2019-01977). The study was performed in accordance with the relevant guidelines and regulations.

## Author contributions

JS and PM designed the study. PM localized electrode positions and resection volumes. SM, MS and SV treated the patients. VD and EB analyzed the data and prepared figures and tables. JS, VD, PM, and SV wrote the manuscript. All authors critically reviewed the manuscript.

## References

1. Ryvlin P, Rheims S. Predicting epilepsy surgery outcome. Curr Opin Neurol. 2016;29(2):182–8.

2. Rosenow F, Luders H. Presurgical evaluation of epilepsy. Brain : a journal of neurology. 2001;124(Pt 9):1683–700.

3. Jobst BC, Bartolomei F, Diehl B, Frauscher B, Kahane P, Minotti L, Sharan A, Tardy N, Worrell G, Gotman J. Intracranial EEG in the 21st Century. Epilepsy Curr. 2020;20(4):180–8.

4. Megevand P, Spinelli L, Genetti M, Brodbeck V, Momjian S, Schaller K, Michel CM, Vulliemoz S, Seeck M. Electric source imaging of interictal activity accurately localises the seizure onset zone. J Neurol Neurosurg Psychiatry. 2014;85(1):38–43.

5. van ‘t Klooster MA, van Klink Nec, Zweiphenning W, Leijten FSS, Zelmann R, Ferrier CH, van Rijen PC, Otte WM, Braun KPJ, Huiskamp GJM, Zijlmans M. Tailoring epilepsy surgery with fast ripples in the intraoperative electrocorticogram. Annals of neurology. 2017;81(5):664–76.

6. Fedele T, van ‘t Klooster M, Burnos S, Zweiphenning W, van Klink N, Leijten F, Zijlmans M, Sarnthein J. Automatic detection of high frequency oscillations during epilepsy surgery predicts seizure outcome. Clinical neurophysiology. 2016;127(9):3066–74.

7. Fedele T, Burnos S, Boran E, Krayenbuhl N, Hilfiker P, Grunwald T, Sarnthein J. Resection of high frequency oscillations predicts seizure outcome in the individual patient. Sci Rep. 2017;7(1):13836.

8. Fedele T, Ramantani G, Sarnthein J. High frequency oscillations as markers of epileptogenic tissue - End of the party? Clinical neurophysiology. 2019;130(5):624–6.

9. Jacobs J, Zijlmans M. HFO to Measure Seizure Propensity and Improve Prognostication in Patients With Epilepsy. Epilepsy Curr. 2020:1535759720957308.

10. Boran E, Sarnthein J, Krayenbuhl N, Ramantani G, Fedele T. High-frequency oscillations in scalp EEG mirror seizure frequency in pediatric focal epilepsy. Sci Rep. 2019;9(1):16560.

11. Liu S, Quach MM, Curry DJ, Ummat M, Seto E, Ince NF. High-frequency oscillations detected in ECoG recordings correlate with cavernous malformation and seizure-free outcome in a child with focal epilepsy: A case report. Epilepsia Open. 2017;2(2):267–72.

12. Akiyama T, McCoy B, Go CY, Ochi A, Elliott IM, Akiyama M, Donner EJ, Weiss SK, Snead OC, 3rd, Rutka JT, Drake JM, Otsubo H. Focal resection of fast ripples on extraoperative intracranial EEG improves seizure outcome in pediatric epilepsy. Epilepsia. 2011;52(10):1802–11.

13. Jacobs J, Zijlmans M, Zelmann R, Chatillon CE, Hall J, Olivier A, Dubeau F, Gotman J. High-frequency electroencephalographic oscillations correlate with outcome of epilepsy surgery. Annals of neurology. 2010;67(2):209–20.

14. Sumsky SL, Santaniello S. Decision Support System for Seizure Onset Zone Localization Based on Channel Ranking and High-Frequency EEG Activity. IEEE J Biomed Health Inform. 2019;23(4):1535–45.

15. Cuello-Oderiz C, von Ellenrieder N, Sankhe R, Olivier A, Hall J, Dubeau F, Gotman J. Value of ictal and interictal epileptiform discharges and high frequency oscillations for delineating the epileptogenic zone in patients with focal cortical dysplasia. Clinical neurophysiology. 2018;129(6):1311–9.

16. Schonberger J, Huber C, Lachner-Piza D, Klotz KA, Dumpelmann M, Schulze-Bonhage A, Jacobs J. Interictal Fast Ripples Are Associated With the Seizure-Generating Lesion in Patients With Dual Pathology. Front Neurol. 2020;11:573975.

17. Cimbalnik J, Kucewicz MT, Worrell G. Interictal high-frequency oscillations in focal human epilepsy. Curr Opin Neurol. 2016;29(2):175–81.

18. Weiss SA, Berry B, Chervoneva I, Waldman Z, Guba J, Bower M, Kucewicz M, Brinkmann B, Kremen V, Khadjevand F, Varatharajah Y, Guragain H, Sharan A, Wu C, Staba R, Engel J, Jr., Sperling M, Worrell G. Visually validated semi-automatic high-frequency oscillation detection aides the delineation of epileptogenic regions during intra-operative electrocorticography. Clinical neurophysiology. 2018;129(10):2089–98.

19. Nariai H, Hussain SA, Bernardo D, Fallah A, Murata KK, Nguyen JC, Rajaraman RR, Rao LM, Matsumoto JH, Lerner JT, Salamon N, Elashoff D, Sankar R, Wu JY. Prospective observational study: Fast ripple localization delineates the epileptogenic zone. Clinical neurophysiology. 2019;130(11):2144–52.

20. Remakanthakurup Sindhu K, Staba R, Lopour BA. Trends in the use of automated algorithms for the detection of high-frequency oscillations associated with human epilepsy. Epilepsia. 2020;61(8):1553–69.

21. Burnos S, Frauscher B, Zelmann R, Haegelen C, Sarnthein J, Gotman J. The morphology of high frequency oscillations (HFO) does not improve delineating the epileptogenic zone. Clinical neurophysiology. 2016;127(4):2140–8.

22. Groppe DM, Bickel S, Dykstra AR, Wang X, Megevand P, Mercier MR, Lado FA, Mehta AD, Honey CJ. iELVis: An open source MATLAB toolbox for localizing and visualizing human intracranial electrode data. J Neurosci Methods. 2017;281:40–8.

23. Frauscher B, von Ellenrieder N, Zelmann R, Rogers C, Nguyen DK, Kahane P, Dubeau F, Gotman J. High-Frequency Oscillations in the Normal Human Brain. Annals of neurology. 2018;84(3):374–85.

24. Jacobs J, Wu JY, Perucca P, Zelmann R, Mader M, Dubeau F, Mathern GW, Schulze-Bonhage A, Gotman J. Removing high-frequency oscillations: A prospective multicenter study on seizure outcome. Neurology. 2018;91(11):e1040–e52.

25. Levesque M, Wang S, Gotman J, Avoli M. High frequency oscillations in epileptic rodents: Are we doing it right? J Neurosci Methods. 2018;299:16–21.

26. Garcia Gracia C, Yardi R, Kattan MW, Nair D, Gupta A, Najm I, Bingaman W, Gonzalez-Martinez J, Jehi L. Seizure freedom score: A new simple method to predict success of epilepsy surgery. Epilepsia. 2015;56(3):359–65.

27. Wu JY, Sankar R, Lerner JT, Matsumoto JH, Vinters HV, Mathern GW. Removing interictal fast ripples on electrocorticography linked with seizure freedom in children. Neurology. 2010;75(19):1686–94.

28. Frauscher B. Localizing the epileptogenic zone. Curr Opin Neurol. 2020;33(2):198–206.

